# Longitudinal Profiling of the Intestinal Microbiome in Children with Cystic Fibrosis Treated with Elexacaftor-Tezacaftor-Ivacaftor

**DOI:** 10.1101/2023.08.11.23293949

**Authors:** Seth A. Reasoner, Rachel Bernard, Adam Waalkes, Kelsi Penewit, Janessa Lewis, Andrew G. Sokolow, Rebekah F. Brown, Kathryn M. Edwards, Stephen J. Salipante, Maria Hadjifrangiskou, Maribeth R. Nicholson

## Abstract

The intestinal microbiome influences growth and disease progression in children with cystic fibrosis (CF). Elexacaftor-tezacaftor-ivacaftor (ELX/TEZ/IVA), the newest pharmaceutical modulator for CF, restores function of the pathogenic mutated CFTR channel. We performed a single-center longitudinal analysis of the effect of ELX/TEZ/IVA on the intestinal microbiome, intestinal inflammation, and clinical parameters in children with CF. Following ELX/TEZ/IVA, children with CF had significant improvements in BMI, ppFEV_1_ and required fewer antibiotics for respiratory infections. Intestinal microbiome diversity increased following ELX/TEZ/IVA coupled with a decrease in the intestinal carriage of *Staphylococcus aureus*, the predominant respiratory pathogen in children with CF. There was a reduced abundance of microbiome-encoded antibiotic-resistance genes. Microbial pathways for aerobic respiration were reduced after ELX/TEZ/IVA. The abundance of microbial acid tolerance genes was reduced, indicating microbial adaptation to increased CFTR function. In all, this study represents the first comprehensive analysis of the intestinal microbiome in children with CF receiving ELX/TEZ/IVA.

**IMPORTANCE:** Cystic fibrosis is an autosomal recessive disease with significant gastrointestinal symptoms in addition to pulmonary complications. Prior work has shown that the intestinal microbiome correlates with health outcomes in CF, particularly in children. Recently approved treatments for CF, CFTR modulators, are anticipated to substantially improve the care of patients with CF and extend their lifespans. Here, we study the intestinal microbiome of children with CF before and after the CFTR modulator, ELX/TEZ/IVA. We identify promising improvements in microbiome diversity, reduced measures of intestinal inflammation, and reduced antibiotic resistance genes. We present specific bacterial taxa and protein groups which change following ELX/TEZ/IVA. These results will inform future mechanistic studies to understand the microbial improvements associated with CFTR modulator treatment. This study demonstrates how the microbiome can change in response to a targeted medication that corrects a genetic disease.

## 1. INTRODUCTION

Cystic fibrosis (CF) is an autosomal recessive disease affecting a total of 40,000 individuals in the United States (1). CF is caused by mutations in the CF transmembrane conductance regulator (*CFTR*) gene resulting in decreased epithelial transport of chloride and bicarbonate ions. These mutations result in mucus obstruction which presents with severe multi-organ dysfunction, principally affecting the airways and gastrointestinal tract (2–5). The gastrointestinal complications include malnutrition, dysmotility, and hepatopancreaticobiliary disease (2). Importantly, nutritional status and intestinal microbiome abnormalities in children with CF have been linked to growth failure, disease progression, and risk of future lung transplantation (6–11).

Multiple studies have demonstrated differences in the intestinal microbiome of patients with CF when compared to healthy controls (8, 12–15). The CF intestinal microbiome is notably inflammatory and associated with higher rates of inflammatory bowel disease (IBD) and colon cancer (16–20). In infants with CF, delayed intestinal microbiome maturation has been shown to influence linear growth and immune programming (8, 21). Among the most striking differences compared to healthy controls is the abundance of Proteobacteria, specifically *Escherichia coli*, and concomitant decrease in Bacteroidetes in infants with CF (8, 12, 22–24). Furthermore, during acute CF pulmonary exacerbations, the intestinal microbiome is distinguishable from periods of respiratory stability (15, 21, 25). Respiratory pathogens, such as *Staphylococcus aureus*, can also be detected in CF stool samples (22, 26).

Over the past decade, small molecule therapies which address the primary defect in the CFTR protein and rescue CFTR function in select genotypes have been developed. These “CFTR modulators” have dramatically changed the trajectory of CF patient care, yielding remarkable improvements in lung function, growth, and projected lifespan (27). The most recent CFTR modulator formulation approved for clinical care, elexacaftor-tezacaftor-ivacaftor (ELX/TEZ/IVA), includes two CFTR correctors (ELX & IVA) and one CFTR potentiator (TEZ) (28). The approval of ELX/TEZ/IVA is anticipated to be the most important advancement in CF therapy since *CFTR* was identified over 30 years ago (27, 28).

Given the progressive nature of CF, initiation of therapy in early childhood is critical to stall disease progression. ELX/TEZ/IVA was approved by the FDA for pediatric patients in 2019 (ages 12 years and older), 2021 (ages 6-11 years old), and 2023 (ages 2-5 years old). The impact of ELX/TEZ/IVA on clinical outcomes and the intestinal microbiome in children with CF requires targeted study. To date, studies examining the effects of CFTR modulators on the intestinal microbiome have been limited by the use of single modulator formulations or small cohort sizes (29–31). Importantly, no studies have characterized the effect of ELX/TEZ/IVA on the intestinal microbiome of children with CF. We therefore undertook a longitudinal study to identify changes in clinical outcomes and the intestinal microbiomes of pediatric patients following the initiation of ELX/TEZ/IVA. To our knowledge, this study represents the first comprehensive study of the highly effective CFTR modulator formulation ELX/TEZ/IVA on the intestinal microbiome of children with CF.

## 2. METHODS

### 2.1 Recruitment of Subjects

Pediatric patients with a diagnosis of CF at Monroe Carell Jr. Children’s Hospital at Vanderbilt (MCJCHV) were recruited beginning in July 2017 with follow-up until October 2022. Patients with the Phe508 CFTR mutation who were deemed eligible for ELX/TEZ/IVA by their pulmonologist were eligible for this study. Patients previously treated with other CFTR modulator regimens were permitted. Patients with pre-existing non-CF gastrointestinal disease were excluded. A total of 39 participants were recruited (Table 1). Informed consent was obtained from parental guardians, and assent was obtained from pediatric subjects in accordance with institutional research ethics guidelines. This study was approved by the MCJCHV Institutional Review Board (IRB # 200396). The lead investigators of this study had no direct role in the patients’ routine medical care. All study data were stored in a Research Electronic Data Capture (REDCap) database per institutional guidelines (32).

**Table 1.** Cohort Baseline Details.

### 2.2 Study Timepoints

Our analysis included stool samples from up to four timepoints per patient: two timepoints before ELX treatment (T1 & T2) and two timepoints after initiating ELX/TEZ/IVA (T3 & T4) (Fig. 1B). For clinical outcomes, data was either analyzed by individual timepoints (T1-T4) or time points were combined for pre-ELX/TEZ/IVA samples (T1 & T2 combined) and post-ELX-TEZ/IVA samples (T3 & T4 combined). Analysis of individual timepoints permitted assessment of additional differences between the 6 to 12-month periods after ELX/TEZ/IVA initiation. For microbiome comparisons, analysis was based on combined pre-ELX/TEZ/IVA samples (T1 & T2) and post-ELX-TEZ/IVA samples (T3 & T4).

**Figure 1.**
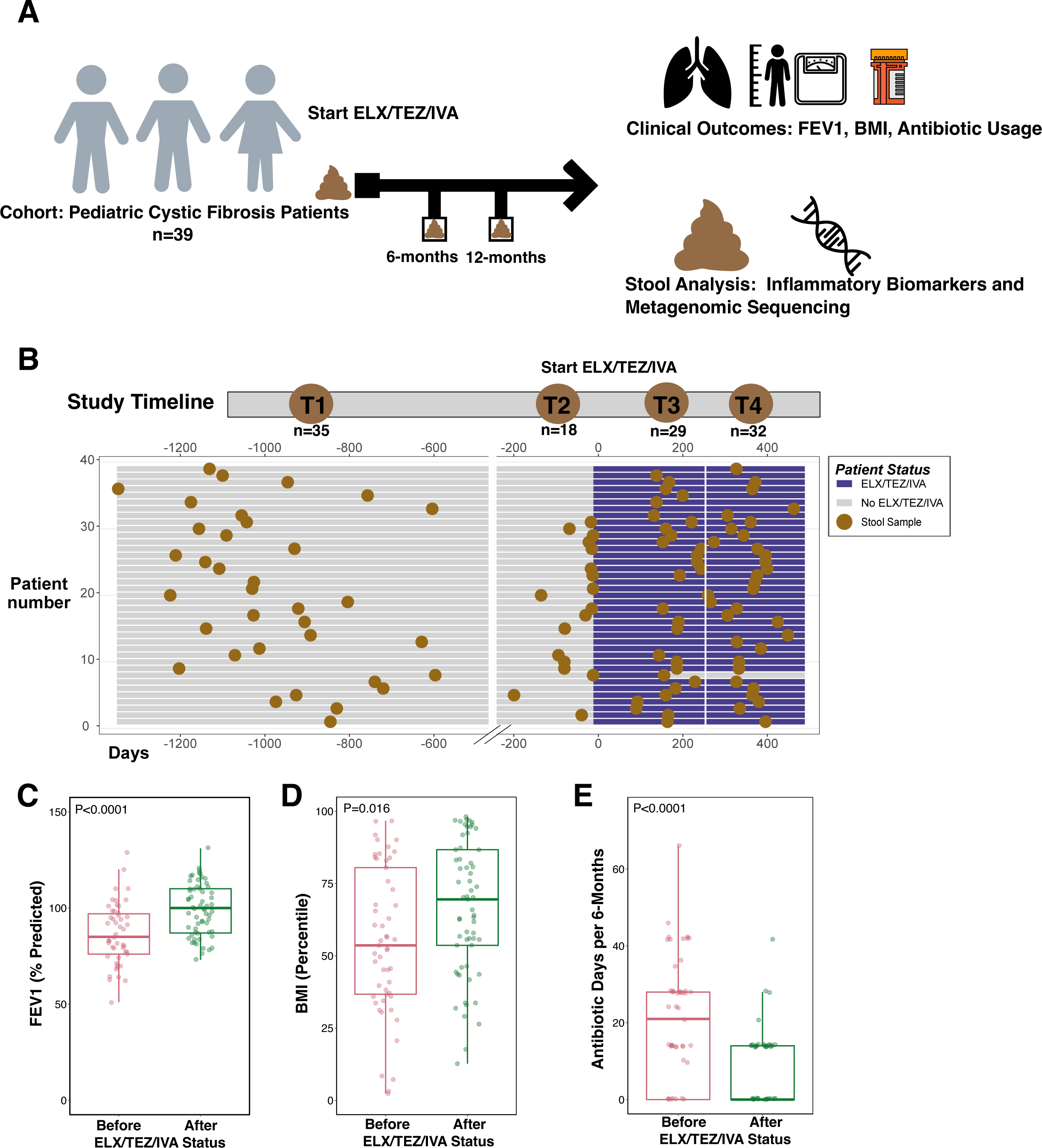
Study Schematic, Timeline, and Clinical Improvement after ELX/TEZ/IVA. **A)** Overview of study procedure and analyses. **B)** Timeline illustrating the four timepoints. Day 0 is depicted as the day of ELX/TEZ/IVA initiation. A total of 114 samples were collected from 39 unique patients, of which 53 samples were before ELX/TEZ/IVA treatment and 61 samples after treatment. **C-E)** Depictions of clinical data before and after ELX/TEZ/IVA; **(C)** ppFEV1, **(D)** BMI percentile, **(E)** antibiotic days per 6-months. Each dot represents the clinical data associated with a stool sample. P values calculated by Wilcoxon rank-sum test.

### 2.3 Stool Sample Collection and DNA Extraction

Stool samples were collected prior to ELX/TEZ/IVA initiation, either through targeted collection as part of this study or from our center’s biobank of CF stool samples (33, 34). Subsequent stool samples were collected at approximately 6- and 12-month intervals after initiation of ELX/TEZ/IVA (Fig. 1A). Fecal samples were collected in sterile collection cups and refrigerated until transport to the laboratory. Patients who were unable to provide a stool sample in clinic were provided with an OMNIgene®•GUT stool collection kit for at home collection and stabilization, returned via overnight shipping, and stored according to manufacturer specifications. Aliquots of stool samples were aseptically aliquoted into cryovials in a laminar flow biosafety cabinet to minimize aerosols and stored at −80°C until processing. Percent predicted forced expiratory volume in one second (ppFEV_1_) values were obtained in the outpatient setting at the time of stool sample collection; the highest spirometry value of three attempts was recorded. Clinical data including body mass index (BMI, percentiles), percentage predicted forced expiratory volume in 1 second (ppFEV_1_), medications, and laboratory values were recorded for visits when a stool sample was collected. Total DNA was extracted from 114 stool samples using QIAamp PowerFecal Pro DNA Kits according to the manufacturer’s instructions. Bead beating for efficient lysis was conducted for 10 minutes. All steps, excluding bead beating and centrifugation, were conducted in a laminar flow biosafety cabinet. No human DNA depletion or enrichment of microbial DNA was performed. DNA yield was estimated by spectrophotometry (NanoDrop™2000c) in parallel with ensuring satisfactory A260/A280 ratio for DNA purity.

### 2.4 Fecal Calprotectin Measurements

Fecal calprotectin was measured using the Calprotectin ELISA Assay Kit (Eagle Biosciences). Duplicate portions of stool (50-100mg) were weighed and processed according to the manufacturer’s instructions. Absorbance was measured with a SpectraMax® i3x (Molecular Devices). Fecal calprotectin (µg/g feces) was calculated using a 7-point standard curve. The assay’s reported normal cut-off is <43.2 μg/g.

### 2.5 Shotgun Metagenomic Sequencing

Sequencing libraries were prepared using Illumina® reagents as described elsewhere (35). Pooled libraries were sequenced on NextSeq2000 to generate 150-bp paired-end reads. An average of 13.9 million reads were produced per sample (range 9.1 to 30.1 million). Sequencing adaptors and low-quality sequences were trimmed with fastq-mcf from ea-utils-1.1.2.779 using default parameters (Supplementary Methods) (36).

### 2.6 Taxonomic and Functional Profiling of Metagenomic Sequence Data

Species abundances were determined with MetaPhlAn4 following read alignment to the MetaPhlan4 database (37). Diversity metrics were calculated with the vegan R package (38) and plotted with ggplot2. A weighted UniFrac distance matrix was constructed with the MetaPhlAn R script “calculate_unifrac”. Functional profiling was conducted with HUMAnN 3.0 with mapping to UniRef90 gene-families and MetaCyc metabolic pathways (39–41). UniRef90 gene-families were reformatted into KEGG orthology (KO) groups using the HUMAnN command “humann_regroup_table”. In total, we identified 722 species, 526 MetaCyc pathways, and 8634 distinct KO groups annotations in the 114 samples. All functional annotations were normalized using the HUMAnN command “humann_renorm_table --units cpm”. Antibiotic resistance genes were identified by ShortBRED (42) with the reference Comprehensive Antibiotic Resistance Database (CARD version 2017) (43). ARG abundance was calculated as reads per kilobase of reference sequence per million sample reads (RPKM).

### 2.7 Microbial Dysbiosis Index

The Microbial Dysbiosis Index (MD-index) was calculated as the log10 of the ratio of the relative abundance of taxa which were previously positively and negatively associated with newly diagnosed pediatric Crohn’s disease (44). Specifically, the numerator includes *Enterobacteriaceae*, *Pasteurellaceae*, *Fusobacteriaceae*, *Neisseriaceae*, *Veillonellaceae*, *Gemellaceae*; the denominator includes *Bacteroidales*, *Clostridiales* (excluding *Veillonellaceae*), *Erysipelotrichaceae*, and *Bifidobacteriaceae*. Higher indices correspond to a greater inflammatory taxonomic profile. The MD-index has been previously applied to stool samples of children with CF (14, 45).

### 2.8 Differential Abundance Testing

We used MaAsLin2 (Microbiome Multivariable Associations with Linear Models, Maaslin2 R package) to identify differentially abundant features (46). We included ELX/TEZ/IVA, age, and recent antibiotic exposure as fixed effects. Subject ID was specified as a random effect due to multiple samples from the same subject. Species, KEGG orthologs, and MetaCyc pathways detected at least 10% of samples were tested (i.e., prevalence = 0.1); no minimum abundance was specified. Abundances were log transformed within the MaAsLin2 function. The general linear “LM” model was used. MaAsLin2 coefficients are equivalent to log2(FoldChange). The Benjamini-Hochberg procedure was used to correct P values, and corrected P values are reported as False Discovery Rates (FDR).

### 2.9 Statistical Analyses

Statistical analyses were conducted using GraphPad Prism 9 and R (version 4.2.1) software. Details of the statistical tests used and the significance thresholds are presented in the figure legends. All box-plot graphs are defined as: center line—median; box limits—upper and lower quartiles; whiskers—1.5× interquartile range.

### 2.10 Reproducibility and Data Availability

The results can be reproduced using raw sequence data that are available on NCBI-Genbank databases under BioProject PRJNA948536 or using processed data in the Supplementary Dataset 1. Bioinformatic code is available within the repository https://github.com/reaset41/CF-GI-Microbiome-ELX-TEZ-IVA.

## 3. RESULTS

### 3.1 Study Cohort and Stool Sample Collection

Stool samples were collected from a total of 39 children with CF (Table 1). The median (IQR) age of participants at the time of ELX/TEZ/IVA initiation was 9.86 years old (8.87, 12.15) and 53.4% were male. Of these, 35.9% (14/39 subjects) had CFTR modulator use (at any time) before ELX/TEZ/IVA. Patients provided an average of 2.92 stool samples with all patients providing at least one stool sample prior to starting ELX/TEZ/IVA (T1 median= 33.6 months before ELX/TEZ/IVA initiation, IQR 28.6, 36.7 months and T2 median=0.64 months before ELX/TEZ/IVA initiation, IQR 0.13, 2.2 months). Subsequent stool samples were collected at approximately 6- and 12-month intervals (T3 median=5.97 months, IQR 5.38, 6.92 months and T4 median=12.39 months, IQR 11.1, 13.1 months). There was a total of 114 stool samples with 53 samples before ELX/TEZ/IVA therapy and 61 samples after ELX/TEZ/IVA treatment (Fig. 1A & S2). Of the 53 samples collected pre-ELX/TEZ/IVA, 8 samples (15%) were collected while the patient was receiving an alternative CFTR modulator (Supplementary Dataset 1).

### 3.2 Clinical Improvement after ELX/TEZ/IVA

To determine our cohort’s clinical response to ELX/TEZ/IVA, we compared clinical metrics from before and after ELX/TEZ/IVA initiation. Both BMI percentile and ppFEV_1_ increased in the timepoints after ELX/TEZ/IVA compared to pre-ELX/TEZ/IVA timepoints (Fig. 1C-D & S1A-B). Median BMI percentile increased from the 54^th^ percentile to the 67^th^ percentile after ELX/TEZ/IVA (Fig. 1D). Likewise, there was a mean change in ppFEV_1_ of 12.3 percentage points (95% CI 6.7-17.8) before and after ELX/TEZ/IVA (Fig. 1C). Between 6- and 12-months after ELX/TEZ/IVA, there was no further significant improvement in either BMI or ppFEV_1_ (Fig. S1A-B, T3 & T4). Consistent with increased BMI, weight percentile increased after ELX/TEZ/IVA (Fig. S1C). Height percentile increased between T2 and both T3 and T4 (Fig. S1D, P=0.013, P=0.026, respectively), though not between T1 and T3 & T4. Total antibiotic days per patient were aggregated and decreased from a median of 22.5 days per 6-months before ELX/TEZ/IVA to 0 antibiotic days per 6-months after ELX/TEZ/IVA (Fig. 1E, P<0.0001).

### 3.3 Microbiome Diversity Increases After ELX/TEZ/IVA

From shotgun metagenomic sequencing on 114 stool samples, we compared measures of microbiome taxonomy and diversity. In contrast to previously published CF microbiome datasets of infants (8, 22), our dataset of older children was dominated by the phylum Firmicutes with minimal Proteobacteria (Fig. 2A and Supplementary Dataset 1). Alpha diversity, as measured by the Shannon index and richness (observed species), significantly increased following ELX/TEZ/IVA (Fig. 2B-C, P=0.021 and P=0.026, respectively). The number of species observed increased from a median (IQR) of 83 (62, 115) species before ELX/TEZ/IVA to 109 (82,125) species after ELX/TEZ/IVA (Fig. 2B). The cumulative increases in alpha diversity were not significantly different when comparing between the four timepoints individually (Fig. S2A-B), and there were no differences in overall community composition (beta-diversity) before and after ELX/TEZ/IVA (Fig. S2C-D).

**Figure 2.**
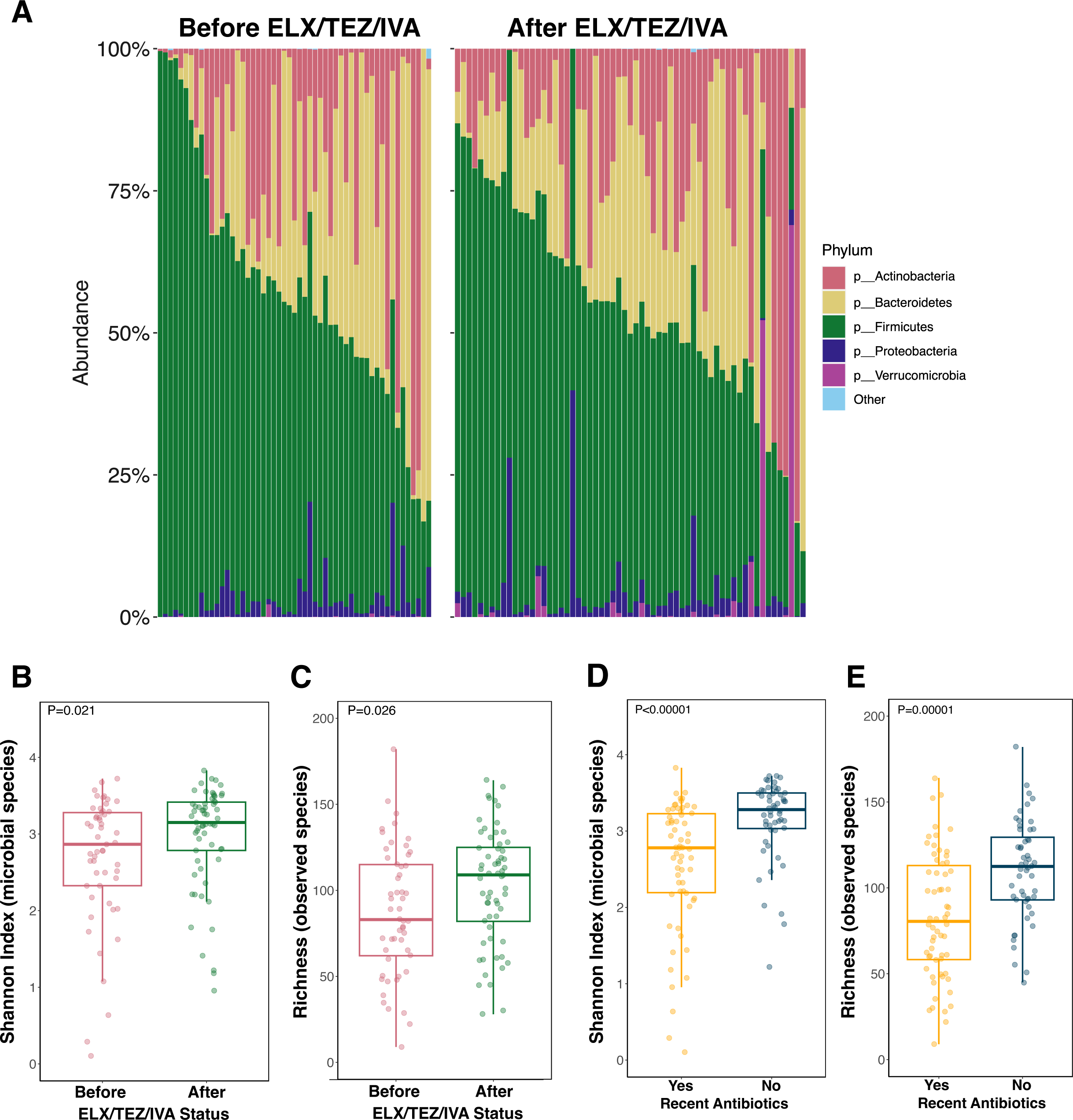
Microbiome Diversity and Intestinal Carriage of Antibiotic Resistance Genes. **A)** Phylum level relative abundance for samples collected before (samples=53) and after (samples=61) ELX/TEZ/IVA. Samples are organized by the relative abundance of the phylum Firmicutes. **B-C)** Alpha diversity before and after ELX/TEZ/IVA. Shannon Index calculated with the R package vegan using species abundance table from Metaphlan4. Microbial richness represents the number of unique species per sample. Each dot represents a stool sample (samples=114). P values calculated by Wilcoxon rank-sum test. **D-E)** Alpha diversity between samples with and without recent antibiotic exposure. Recent antibiotic exposure categorized as any systemic antibiotic within the past 6-months. Shannon Index calculated with the R package vegan using species abundance table from MetaPhlAn4. Microbial richness represents the number of unique species per sample. Each dot represents a stool sample (samples=114). P values calculated by Wilcoxon rank-sum test.

To determine the effect of prior CFTR modulator treatment besides ELX/TEZ/IVA, we compared diversity metrics between modulator naïve samples (samples=45) and samples collected while subjects were receiving another CFTR modulator (samples=8) before ELX/TEZ/IVA. Samples collected while subjects were receiving another CFTR modulator had similar diversity to modulator naïve samples (Fig. S3A-B). Following ELX/TEZ/IVA, samples from subjects who had previously received another modulator (samples=20) had similar improvements in microbiome diversity to samples from subjects who had not previously received another modulator (samples=41), indicating that the residual effects of another modulator did not influence microbiome diversity improvement on ELX/TEZ/IVA (Fig. S3C-D).

To determine the influence of antibiotic exposure on microbiome diversity measures regardless of ELX/TEZ/IVA status, we categorized patient stool samples as having received antibiotics within the six-months prior to stool sample collection (samples=62) or no antibiotic receipt (samples=52). Recent antibiotic exposure significantly impacted alpha diversity measures reducing Shannon diversity and richness (Fig. 2D-E). Likewise, population structure (beta-diversity) was significantly different between samples with and without recent antibiotic exposure (Fig. S2E, permutational multivariate analysis of variance [PERMANOVA]=0.003).

### 3.4 Microbiome Encoded Antibiotic Resistance Genes Decrease After ELX/TEZ/IVA

As discussed above, patients required far fewer antibiotics after starting ELX/TEZ/IVA (Fig. 1E & S4A). Since the intestinal microbiome is a reservoir of antibiotic resistant genes (ARGs) (47), we compared the number and type of intestinal ARGs before and after ELX/TEZ/IVA initiation. Across the 114 samples, we detected a total of 309 unique ARGs. The median number of unique ARGs nominally decreased from a median 86 ARGs (IQR 40, 96) before ELX/TEZ/IVA to a median 68 ARGs (IQR 38, 90) after ELX/TEZ/IVA (Fig. S4B, P=0.31). ARG abundance decreased from a median of 1180 RPKM (IQR 815.4, 1761.1) to 829 RPKM (IQR 460.5, 1299.0) (Fig. 3A, P=0.0097) though this effect was likely mediated by reduced antibiotic use.

**Figure 3.**
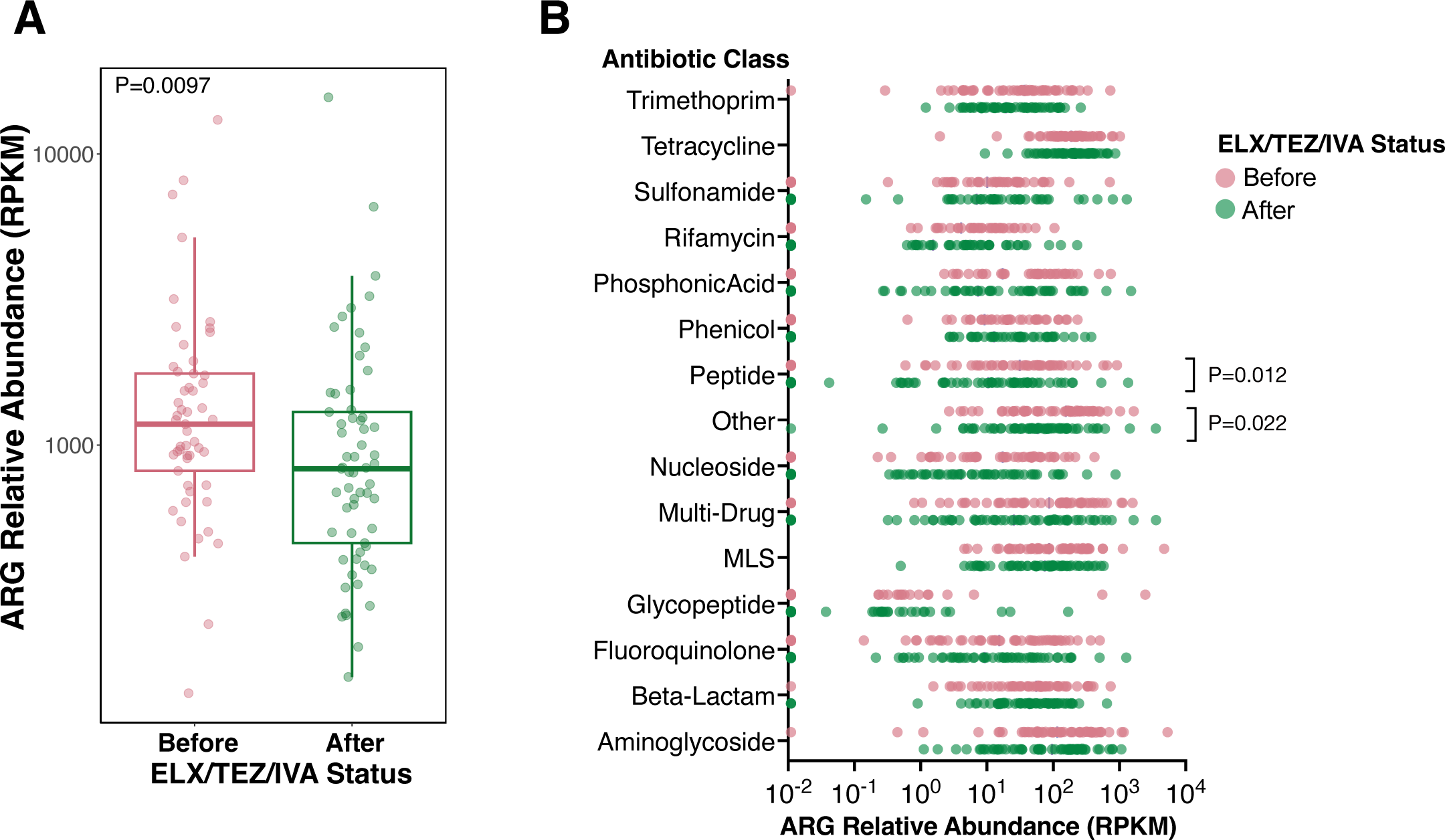
**A)** Antibiotic resistance gene (ARG) abundance (RPKM) before and after ELX/TEZ/IVA. ARGs were profiled using ShortBRED and the Comprehensive Antibiotic Resistance Database (CARD). Each dot represents a stool sample (samples=114). P values calculated by Wilcoxon rank-sum test. **B)** ARG abundance (RPKM) by class of antibiotic to which they confer resistance. Abundance values of zero are plotted on the vertical axis. P values calculated by Wilcoxon signed-rank test.

Next, we aggregated the relative abundance of ARGs by antibiotic class to which they confer resistance (Fig. 3B). The abundance of ARGs conferring resistance to peptide antibiotics and ungrouped antibiotics (“Other”) significantly decreased after ELX/TEZ/IVA (Fig. 3B & S4C-E, P=0.012, P=0.022 respectively). The most prevalent antibiotics within the peptide ARG class included *arnA* (samples=72/114), *yojI* (samples=71/114), *pmrF* (samples=74/114) and *pmrC* (samples=71/114). *arnA, pmrF* and *pmrC* modify Lipid A on bacterial cells to repel cationic peptide antibiotics, whereas *yojI* is a peptide efflux pump. The most prevalent ARGs within the “Other” group were *fabI* (samples=84/114), *gadE* (samples=78/114), and EF-Tu mutations (samples=75/114). These ARGs or mutations confer resistance to isoniazid/disinfecting agents (*fabI*), acid resistance and efflux pump (*gadE*), and elfamycin (EF-Tu). Despite being the most prevalent, none of these ARGs were independently altered in abundance following ELX/TEZ/IVA (Fig. S4D & S4F).

### 3.5 Alterations to Specific Bacterial Taxa Following ELX/TEZ/IVA

To determine whether specific bacterial taxa change following ELX/TEZ/IVA treatment, we used MaAsLin2 (Microbiome Multivariable Associations with Linear Models) to identify differentially abundant microbial taxa (46). No phyla were differentially abundant following ELX/TEZ/IVA (Fig. 2A & Table S4). At a granular taxonomic level, seventeen species were differentially abundant between samples before and after ELX/TEZ/IVA (FDR<0.1, Fig. 4A and Table S3), with eleven species increasing in abundance after ELX/TEZ/IVA, and six species decreasing. The three species with the lowest FDR and, therefore, highest reliability were *Butyricicoccus* SGB14985, *S. aureus,* and *Roseburia faecis* (Fig. 4A-B). *Butyricicoccus* SGB14985, an uncultivated species, was decreased in abundance following ELX/TEZ/IVA (fold change 0.16, FDR=0.022). *Roseburia faecis* was increased in abundance following ELX/TEZ/IVA (fold change 7.35, FDR=0.025) and negatively correlated with recent antibiotic exposure (Fig. S5A, fold change 0.20, FDR=0.086). The intestinal tract has immunological cross-talk between the separate but similarly structured mucosal environment of the lung, deemed the gut-lung axis (20). Since the intestinal tract can harbor similar respiratory pathogens in patients with CF (15, 48, 49), we performed focused analysis of CF pathogens. *S. aureus*, the predominant respiratory pathogen in children with CF (50), was decreased in abundance after ELX/TEZ/IVA (Fig. 4A-B, fold change 0.40, FDR=0.042). Other members of the respiratory microbiota which are frequently detected in the intestines (14, 15)—specifically the genera *Haemophilus*, *Prevotella*, *Streptococcus*, *Veillonella*—did not significantly change after ELX/TEZ/IVA initiation (Fig. S5B). *Pseudomonas aeruginosa*, an additional respiratory pathogen of importance in CF, was detected in only two of the 114 stool samples. Furthermore, fungal taxa were only identified in 8/114 samples (Supplementary Dataset 1).

**Figure 4.**
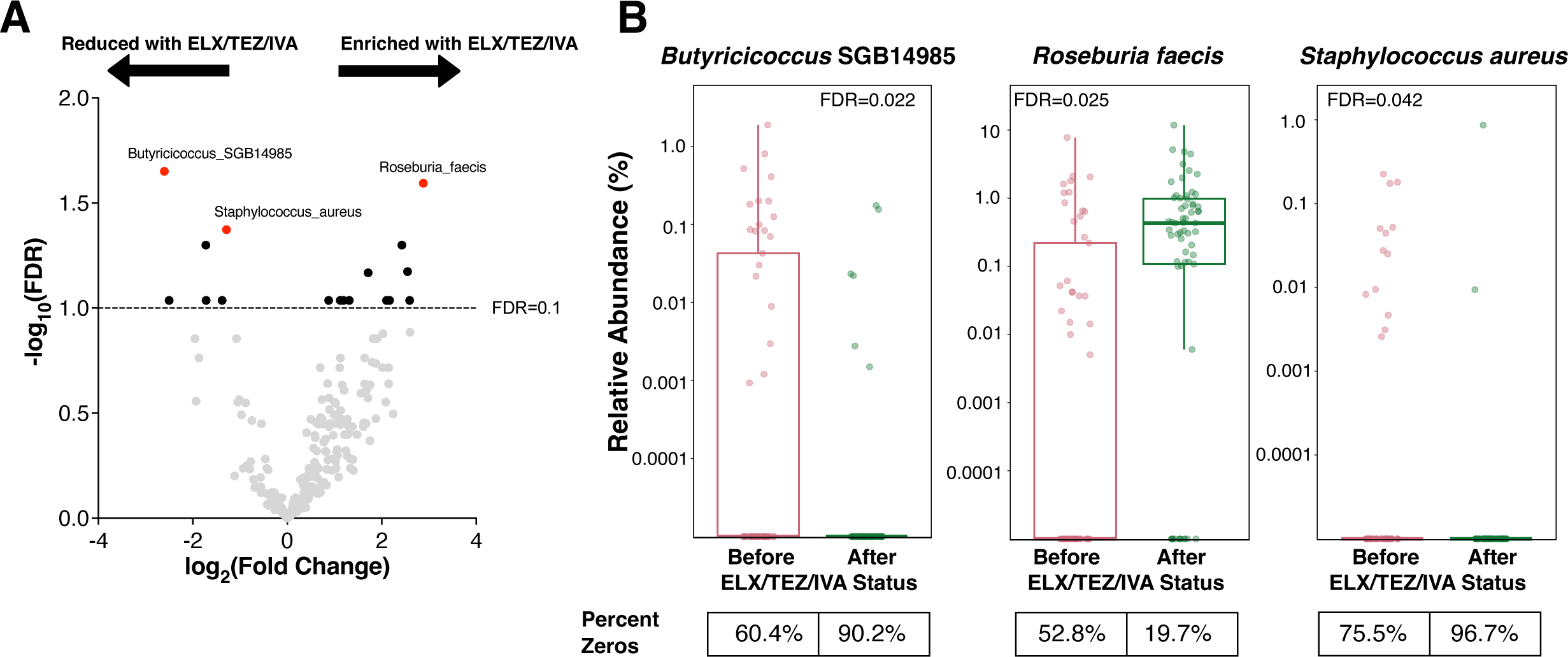
Specific Microbial Taxonomic Changes after ELX/TEZ/IVA. **A)** Differentially abundant species before and after ELX/TEZ/IVA. The results are depicted with significance (−log10 of the FDR) (Y axis) vs. log2(FoldChange). MaAsLin2 multivariable association modeling was implemented in R, using ELX/TEZ/IVA status, age, and recent antibiotics as fixed effects in the model. Participant ID was used as a random effect. Horizontal dashed line depicts FDR=0.1. Species reaching statistical significance (FDR≤0.10) are highlighted in solid colors whereas other species are gray. **B)** Species of interest from differential abundance testing before and after ELX/TEZ/IVA. Vertical axis is log10 transformed. MaAsLin2 FDR is depicted. Zero values are plotted on the horizontal axes. The percentage of samples for which the relative abundance was zero is depicted below the graph.

### 3.6 Intestinal Inflammation Decreases After ELX/TEZ/IVA

Fecal calprotectin was measured across the four timepoints and markedly decreased following ELX/TEZ/IVA (Fig. 5A & S5A). Overall, mean fecal calprotectin decreased from 109 μg/g before ELX/TEZ/IVA to 44.8 μg/g for a mean decrease of 64.2 μg/g (95% CI −16.1, −112.3, Fig. 5A). We then computed the microbial dysbiosis index (MD-index), a ratio of bacterial taxa positively and negatively associated with newly diagnosed pediatric Crohn’s disease, to assess the relationship between intestinal inflammation and specific microbial signatures (44). We noted a nominal reduction in the MD-index following ELX/TEZ/IVA (Fig. 5B, P=0.068), which was less pronounced when comparing samples from patients with recent antibiotic exposure and those without (Fig. 5C, P=0.77).

**Figure 5.**
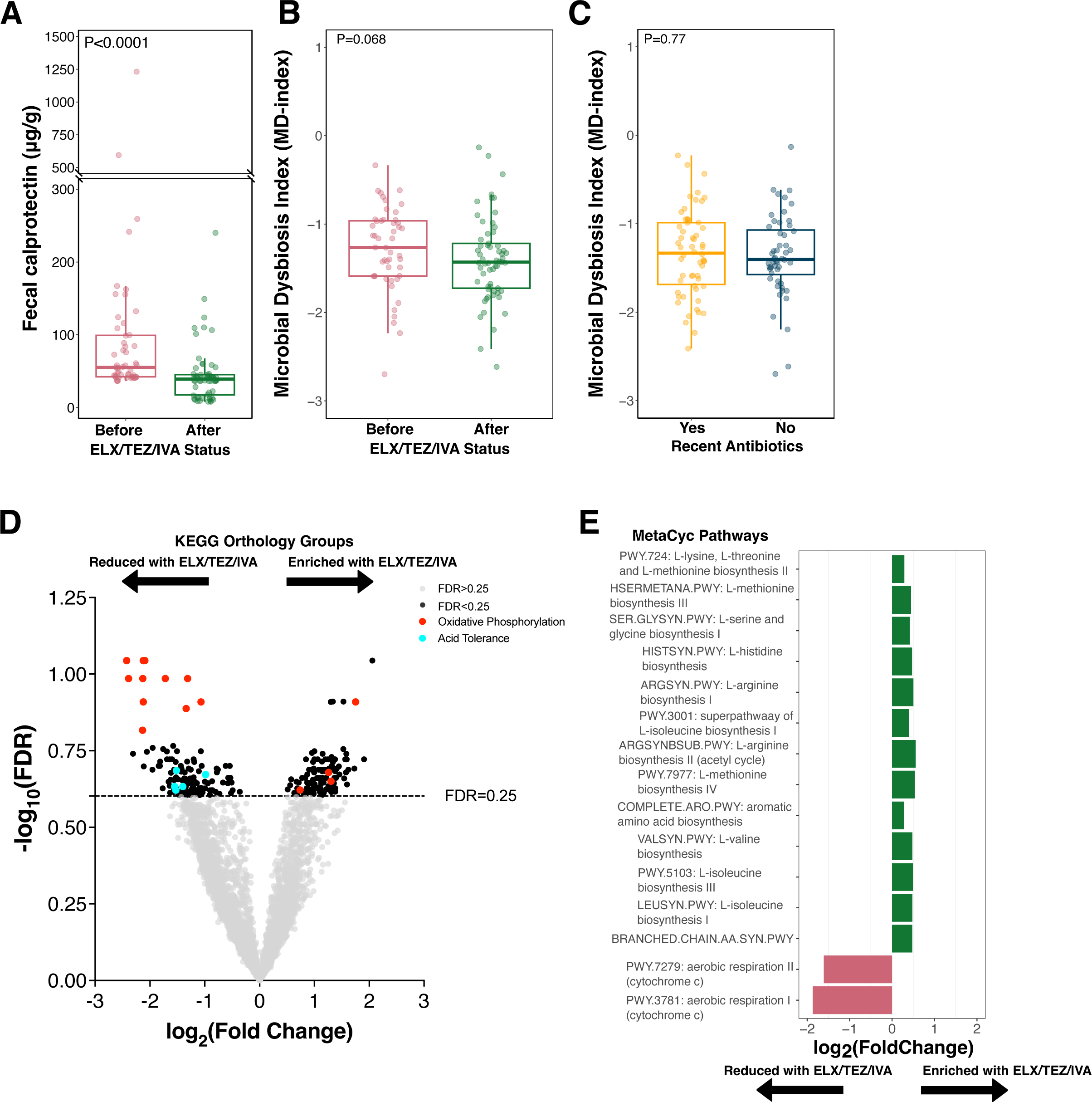
Intestinal Inflammation Decreases after ELX/TEZ/IVA and Microbiome Functional Changes. **A)** Fecal calprotectin before and after ELX/TEZ/IVA. Each dot represents a stool sample (samples=114). P values calculated by Wilcoxon rank-sum test. **B-C)** Microbial dysbiosis index (MD-INDEX) before and after ELX/TEZ/IVA **(B)** or between samples with and without recent antibiotic exposure **(C)**. MD-index calculated as the log10 ratio of species positively/negatively correlated with new onset pediatric Crohn’s disease (44). Each dot represents a stool sample (samples=114). P values calculated by Wilcoxon rank-sum test. **D)** Differentially abundant KEGG orthologs following MaAsLin2 multivariable association modeling, using ELX/TEZ/IVA status, age, and recent antibiotics as fixed effects in the model. Participant ID was used as a random effect. Horizontal dashed line depicts FDR=0.25. **E)** Subset of differentially abundant MetaCyc pathways following MaAsLin2 multivariable association modeling, using ELX/TEZ/IVA status, age, and recent antibiotics as fixed effects in the model. Only pathways with FDR<0.25 are depicted.

### 3.7 Microbiome Functional Changes Reveal Specific Disease-Relevant Changes

From the 114 stool samples, alignment of sequencing reads to known microbial proteins and pathways identified 8634 KEGG orthology (KO) protein groups and 526 MetaCyc metabolic pathways, consistent with prior studies of the intestinal microbiota (51). There were 249 KO groups differentially abundant with respect to ELX/TEZ/IVA use (MaAsLin2 FDR<0.25, Fig. 5D & Table S6). Strikingly, a common thread among differentially abundant KO groups was a decreased abundance of genes encoding oxidative phosphorylation functions following ELX/TEZ/IVA (Fig. 5D). Seven of the top 10 most differentially abundant KO groups were components of the electron transport chain (Fig. 5D & S5B and Table S7) and included NADH-ubiquinone oxidoreductases (e.g., K03883, K03881, K03884, & K03880), F-type ATPase subunits (e.g., K02125), and cytochrome c oxidase subunits (e.g., K02262, K02261, & K02256). The taxa contributing to oxidative phosphorylation KO groups were not consistently annotated by HUMAnN (Fig. S6C and Table S10). This indicates that a specific taxonomic signature was not responsible for the reduced abundance of oxidative phosphorylation KO groups (Fig. S6C). These results were corroborated by MaAsLin2 differential abundance testing of MetaCyc pathways. Sixty-four MetaCyc pathways were differentially abundant with respect to ELX/TEZ/IVA use (MaAsLin2 FDR<0.25, Fig. 5E & S7, Table S9). Two of these pathways corresponded to aerobic respiration (PWY-3781 and PWY-7279). Differential abundance of anaerobic energy metabolism pathways showed discordant results with one pathway increased in abundance following ELX/TEZ/IVA (PWY-7383), and two pathways decreased in abundance (PWY-7389 and PWY-7384) following ELX/TEZ/IVA (Fig. S7 and Table S9). Thirteen amino acid biosynthesis pathways were universally prevalent in all stool samples (114/114) and significantly increased in abundance following ELX/TEZ/IVA (Fig. 5E and Table S9, FDR<0.25). This indicates increased biosynthetic capacity of the intestinal microbiome following ELX/TEZ/IVA.

Furthermore, multiple pathways related to nucleobase and nucleotide metabolism were differentially abundant following ELX/TEZ/IVA. Pyrimidine deoxyribonucleotide biosynthesis pathways were reduced in abundance following ELX/TEZ/IVA (PWY-7184 & PWY-6545) whereas a pyrimidine ribonucleotides biosynthesis pathway was increased in abundance (PWY0-162, Fig.S7 and Table S9). While these general pyrimidine deoxyribonucleotide biosynthesis pathways were reduced in abundance, specific pathways for thiamine biosynthesis (PWY-7357) and two uridine-monophosphate biosynthesis pathways (PWY-7790 & PWY−7791) were significantly increased in abundance following ELX/TEZ/IVA. An anaerobic pathway for purine degradation was increased in abundance following ELX/TEZ/IVA (P164-PWY). Three pathways related to allantoin degradation were significantly reduced in abundance following ELX/TEZ/IVA (PWY-5692, PWY0-41 & URDEGR-PWY, Fig. S7 and Table S9).

Another intriguing observation was the reduced abundance of KO groups involved in acid tolerance and transport following ELX/TEZ/IVA (Fig. 5D). KO groups involved in organic acid transport (e.g., K03290 & K23016) and proton-symporters (e.g., K11102 & K03459) were reduced in abundance following ELX/TEZ/IVA. Likewise, an acidity-responsive transcriptional regulator (K03765, *cadC*) was decreased in abundance following ELX/TEZ/IVA (Fig. 5D and Table S7, fold change 0.42, FDR=0.27).

## 4. DISCUSSION

ELX/TEZ/IVA has changed the trajectory of CF patient care and dramatically improved clinical outcomes. Nutritional status and components of the intestinal microbiome are strong predictors of future clinical outcomes, particularly in children with CF (6–8). We therefore sought to determine the effects of ELX/TEZ/IVA on intestinal inflammation and the intestinal microbiome in children with CF. Herein, we present a comprehensive longitudinal characterization of the intestinal microbiome in children with CF treated with ELX/TEZ/IVA. We identified widespread changes to the intestinal microbiome after ELX/TEZ/IVA including taxonomic composition, reduced carriage of ARGs, and altered microbiota metabolic functions.

Although randomized controlled trials demonstrated that ELX/TEZ/IVA was safe and effective, there are limited post-approval clinical data, particularly in children over two years old (52–56). BMI and ppFEV_1_ are useful markers of clinical response to CFTR modulator treatment in children (57). Our cohort demonstrated significant improvement in ppFEV_1_ and BMI percentile within six-months of starting ELX/TEZ/IVA, with no additional differences at later time points (Fig. S1A-B). This is consistent with prior clinical trial and real-world data showing the greatest impact on nutritional status and anthropometric parameters immediately after ELX/TEX/IVA therapy initiation with subsequent plateau without regression in children (52, 56, 58). Prior baseline BMI, particularly underweight status, has been predicted to be a major determinant of increase in weight gain in patients with CF treated with ELX/TEZ/IVA (59). Furthermore, between the timepoint immediately before ELX/TEZ/IVA initiation (T2) and post-ELX/TEZ/IVA timepoints (T3 & T4), there was a significant increase in height percentile (Fig. S1D). This suggests that CFTR modulator therapy may improve linear growth in children with CF. In sum, these results support the clinical responsiveness of children with CF to ELX/TEZ/IVA.

The gastrointestinal microbiome influences whole-body physiology. CF intestinal microbiome abnormalities begin in infancy, diverging from healthy controls soon after birth (8, 14, 15, 21–23). Intriguingly, the microbiome of patients with CF resembles that of patients with IBD, a group with a characteristically perturbed intestinal microbiome (12, 45, 60). Considerable prior research has characterized the development of the CF intestinal microbiome in infants (8, 13–15, 21–23). Less information exists about the CF intestinal microbiome in children and adolescents (13, 24, 26, 61–65). A consistent finding is that high levels of the phylum Proteobacteria, specifically the species *Escherichia coli*, define the infant CF intestinal microbiome (12, 22). The abundance of Proteobacteria decreases with age in young children with CF (14, 22). In our dataset of older children with CF, Proteobacteria comprised a minority of the identified bacteria (Fig. 2A). In turn, the relative abundance of the phylum Firmicutes is increased in our cohort, likely representing a more stable “adult-like” microbiome.

Direct comparison of our results to studies containing healthy controls is difficult given different sampling and sequencing methodologies. Notably, although our study identified subtle changes in the intestinal microbiome after initiation of ELX/TEZ/IVA in children with CF, the post-ELX/TEZ/IVA intestinal microbiome remains significantly altered from what has been described in healthy children. Compared to healthy children, the intestinal microbiome of children with CF is delayed in development and exhibits decreased microbial diversity relative to healthy controls throughout childhood and adolescence (22, 24, 63). Among the most consistent differences between healthy controls and patients with CF is reduced abundance of the phylum Bacteroidetes in patients with CF (13, 20, 61, 63, 66). In our dataset, Bacteroidetes remained depleted post-ELX/TEZ/IVA (Fig. 2A and Table S4). Likewise, Shannon diversity remained below that of similarly-aged healthy controls (24, 63), as discussed henceforth.

In our cohort, alpha diversity, as measured by the Shannon index and richness (observed species), significantly increased following ELX/TEZ/IVA (Fig. 2B-C). These results demonstrate modest but statistically significant differences in an older cohort of children with CF, whose microbiome resembles that of more of a stable “adult-like” microbiome. The difference in the median Shannon diversity was increased 0.29 after ELX/TEZ/IVA (Fig. 2B). In contrast, a similarly aged cohort of children with CF exhibited a median ∼1.0 reduced Shannon index compared to healthy controls in the same study (24). Thus, while our cohort exhibited significant increases in Shannon diversity, it is unlikely that this magnitude of increase restores the diversity to that of similarly aged healthy controls. This is consistent with the notion that “adult-like” intestinal microbiota are remarkably stable and resilient to intervention (67–69). As CFTR modulator formulations are approved for younger children, particularly infants for whom the microbiome is still developing, studying the effects of CFTR modulators on the developing intestinal microbiome will be an important research endeavor and may show more substantial differences.

In children with CF, markers of intestinal inflammation correlate with growth failure (11, 24). Moreover, intestinal inflammation in patients with CF is linked to higher rates of IBD and colorectal cancer in patients with CF (16, 17, 70). Fecal calprotectin, a laboratory marker of intestinal inflammation (71–73), significantly decreased following ELX/TEZ/IVA (Fig. 5A), corroborating prior reports in the PROMISE and RECOVER cohorts (74, 75). Using a dysbiosis index of bacterial taxa correlated with pediatric Crohn’s disease (44), we identified that this dysbiosis index nominally decreased following ELX/TEZ/IVA initiation (Fig. 5B). The decrease in the median MD-index was 0.166 before and after ELX/TEZ/IVA. The median post-ELX/TEZ/IVA MD-index was lower (i.e. reduced inflammatory taxa) than that of healthy controls in the original publication describing the MD-index (44). This suggests that the magnitude of decrease in the MD-index in our study may be biologically meaningful. Furthermore, the difference between pre- and post-ELX/TEZ/IVA (Fig. 5B) was more pronounced than in samples with and without recent antibiotic exposure (Fig. 5C). This suggests that this effect may be specific to ELX/TEZ/IVA treatment, and not due to reduced antibiotic use. At the species level, the butyrate-producing species *Roseburia faecis* was significantly enriched following ELX/TEZ/IVA (Fig. 4A-B). Prior studies have identified *R. faecis* as reduced in abundance in patients with CF (20, 76–78). In parallel, we detected reduced abundance of genes involved in oxygen-dependent metabolism following ELX/TEZ/IVA (Fig. 5D-E). Intestinal inflammation is characterized by a shift towards oxygen-dependent microbiota metabolism, perpetuating a cycle of inflammatory damage (79–81). From multiple lines of evidence, our results suggest that ELX/TEZ/IVA reduces intestinal inflammation in children with CF.

Respiratory infections require frequent antibiotics in patients with CF. Some respiratory pathogens also colonize the intestinal tract and temporally correlate with respiratory colonization (15, 48, 49). *S. aureus* is among the first pathogens to colonize the respiratory tract and cause infections in children with CF (50). We detected reduced intestinal abundance of *S. aureus* following ELX/TEZ/IVA (Fig. 4A-B). Antibiotic exposure in patients with CF has been associated with increased intestinal carriage of antibiotic resistant bacteria compared to healthy controls which are a poor prognostic factor (82–84). Following ELX/TEZ/IVA, patients in our cohort required far fewer antibiotics (Fig. 1E). In turn, we detected reduced intestinal abundance of ARGs (Fig. 2E). These results indicate disease-relevant taxonomic changes to the intestinal microbiome following ELX/TEZ/IVA, as well as reduced ARGs.

The CFTR channel permits transepithelial movement of bicarbonate (HCO3^-^) and chloride (Cl^-^) (5). Intestinal pH is lower in patients with CF due to the lack of neutralizing bicarbonate (85) yet increases with CFTR modulator treatment, consistent with increased CFTR activity in the intestinal tract (86). We observed a reduced abundance of microbial protein groups associated with acid tolerance (Fig. 5D), consistent with microbial adaptation to increased CFTR channel function in the intestines. These results display physiologically intuitive functional changes to the microbiome post-ELX/TEZ/IVA. As CFTR modulators become the mainstay of treatment for CF, further research will be necessary to mechanistically describe the effects of CFTR modulators on gastrointestinal function.

Our study is strengthened by longitudinal sampling over a five-year period combined with the real-world use of ELX/TEZ/IVA. Using shotgun metagenomic sequencing, we comprehensively compared microbiota differences before and after ELX/TEZ/IVA. Sampling of the intestinal microbiome over extended periods will be important to uncover additional long-term improvements to the microbiome. Similarly, further research is necessary to delineate the effects of ELX/TEX/IVA on the interconnected respiratory and intestinal microbiomes. Although our study was limited by the single-center focus, our cohort mirrors the overall CF pediatric population in clinical and demographic factors (Table 1). Our study lacks untreated CF controls or healthy controls, which would allow for additional comparisons.

In summary, our results indicate that the CFTR modulator ELX/TEZ/IVA alters the intestinal microbiome in children with CF. We identified taxonomic and functional changes to the intestinal microbiome that represent improvements to CF intestinal microbiome structure and function. Our results also support that CFTR modulators reduce intestinal inflammation in children with CF.

## AUTHOR CONTRIBUTIONS (CRediT author statement)

SAR: Investigation, Methodology, Data Curation, Formal analysis, Writing - Original Draft

RB: Conceptualization, Funding Acquisition, Investigation, Data Curation, Writing - Original Draft

AW: Investigation, Software

KP: Investigation JS: Investigation

AGS: Supervision, Writing - Review & Editing

RFB: Supervision, Writing - Review & Editing

KME: Supervision, Writing - Review & Editing

SJS: Investigation, Data Curation, Software, Writing - Review & Editing

MH: Supervision, Writing - Review & Editing

MRN: Conceptualization, Funding Acquisition, Investigation, Supervision, Writing - Original Draft

## Data Availability

All sequence data derived from this work are publicly available in NCBI-Genbank databases under Bioproject PRJNA948536. All processed data and code used for bioinformatic analysis is available within the repository: https://github.com/reaset41/CF-GI-Microbiome-ELX-TEZ-IVA.

## ACKNOWLEDGEMENTS AND FUNDING

We would like to express our sincere appreciation to the patients and their families who enthusiastically participated in this study. We would like to thank our advanced practitioner and nursing colleagues from the Pediatric Pulmonology clinic at the Monroe Carrell Jr. Children’s Hospital at Vanderbilt for their collegiality and assistance with this study. This work was supported, in part, by National Institutes of Health (NIH) grants T32DK007673 (RB), K23AI156132 (MRN), F30AI169748 (SAR), T32GM007347 (SAR), and P30DK089507 (SJS); the Cystic Fibrosis Foundation (SINGH19R0 to SJS) and the Thrasher Research Fund (RB and MRN).

## CONFLICTS OF INTERESTS

All authors declare that they have no conflicts of interest relevant to this work.

## DATA AVAILABILITY

All sequence data derived from this work are publicly available in NCBI-Genbank databases under BioProject PRJNA948536. All processed data and code used for bioinformatic analysis is available within the repository: https://github.com/reaset41/CF-GI-Microbiome-ELX-TEZ-IVA.

---

Supplementary Dataset 1. Compiled Metadata and Metagenomic Results

## FIGURE LEGENDS

**Supplementary Figure S1.**
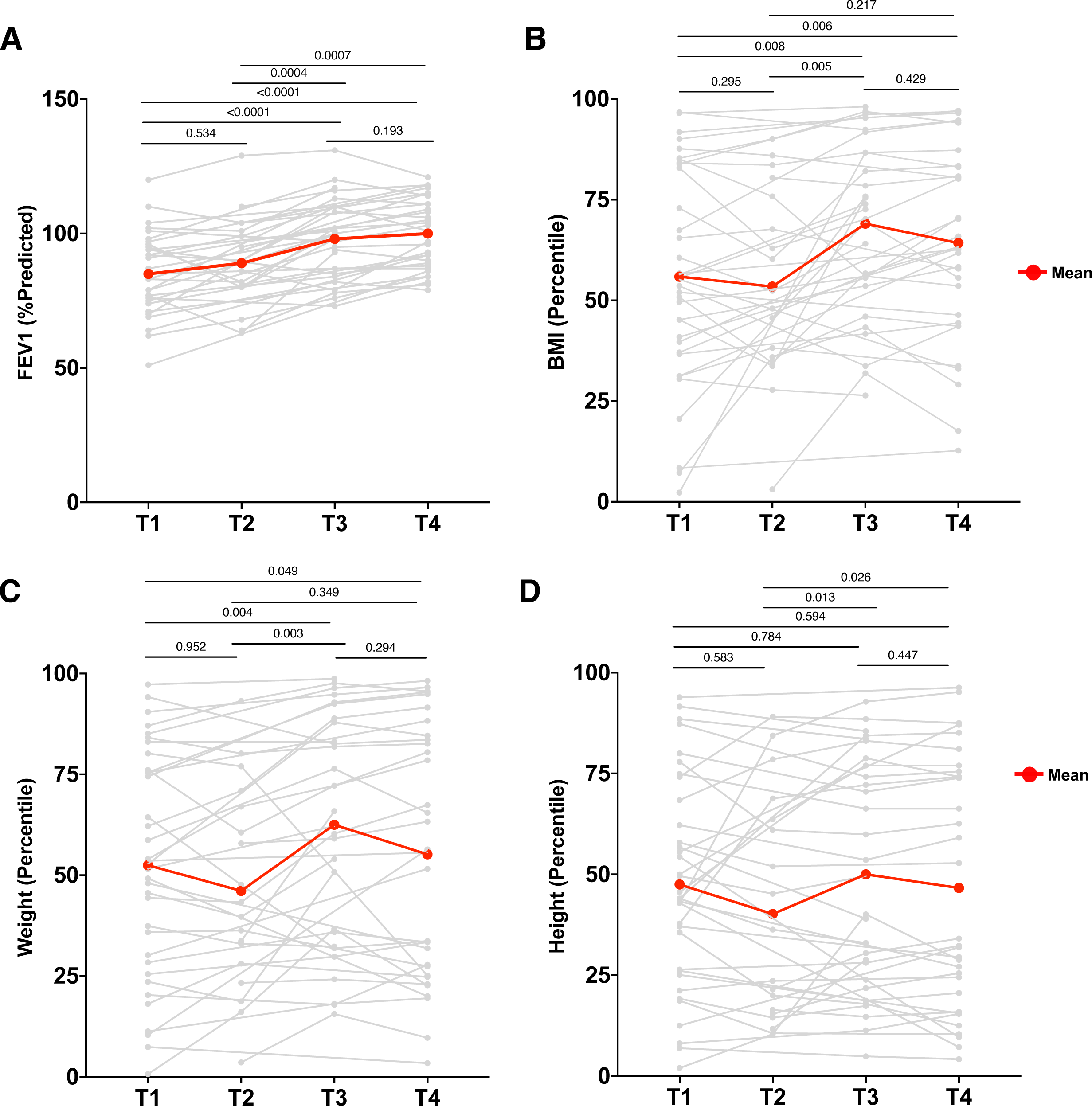
**A-D)** Clinical metadata vs. timepoint; **(A)** ppFEV1, **(B)** BMI percentile, **(C)** weight percentile, and **(D)** height percentile. Each dot represents the clinical data associated with a stool sample. Red line indicates the mean, and gray lines represent individual patients. P values calculated by Wilcoxon signed-rank test.

**Supplementary Figure S2.**
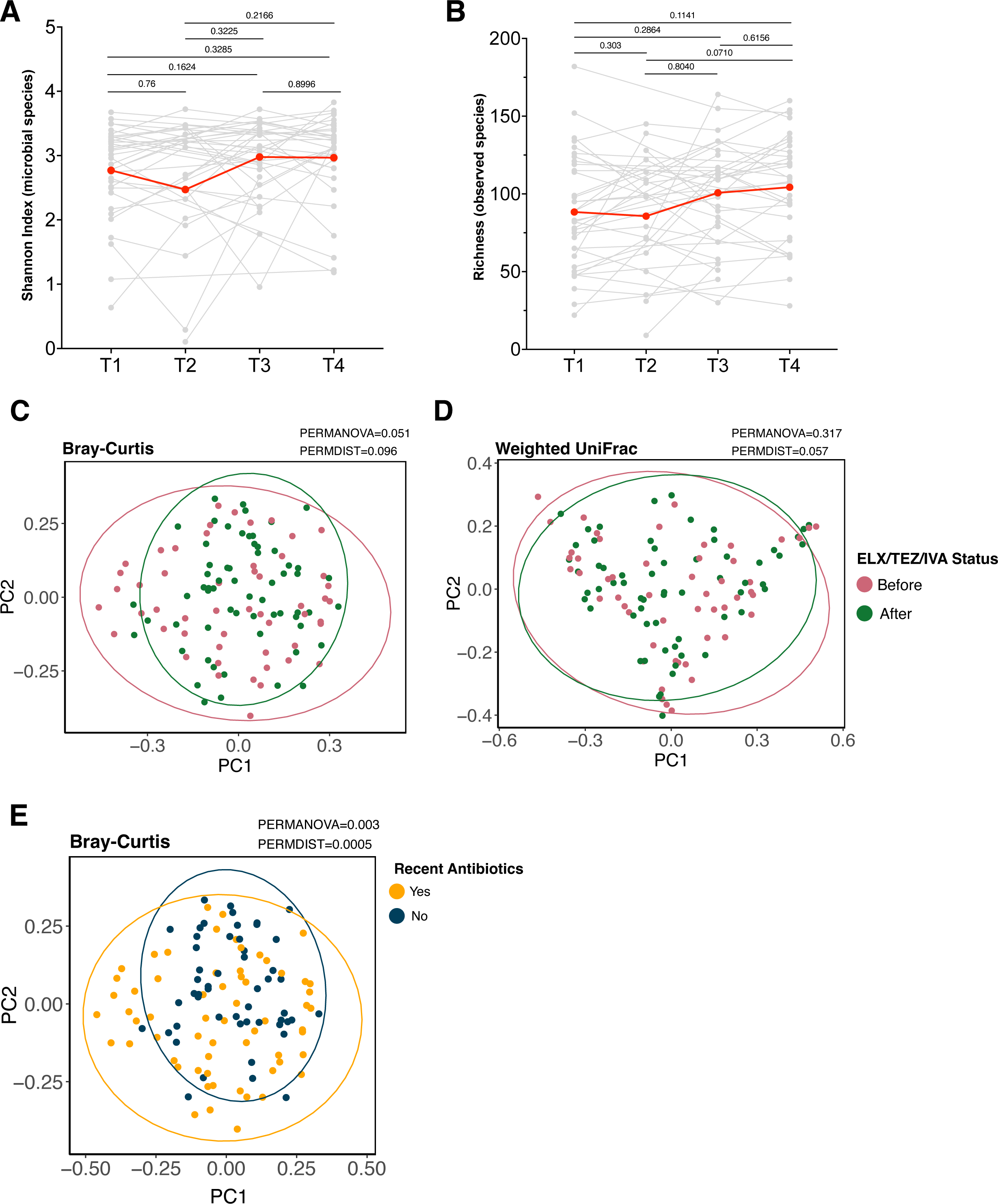
**A-B)** Alpha diversity vs. timepoint. Shannon Index (**A**) calculated with the R package vegan using species abundance table from Metaphlan4. Microbial richness (**B**) represents the number of unique species per sample. Each dot represents a stool sample (samples=114). P values calculated by Wilcoxon signed-rank test. **C)** Principal component analysis of Bray-Curtis distances before and after ELX/TEZ/IVA. Bray-Curtis distance matrix computed with vegan. PERMANOVA and PERMDISP computed by the vegan package with the functions adonis2 and betadisp, respectively. Ellipse depicts the 95% confidence level. **D)** Principal component analysis of weighted UniFrac distances before and after ELX/TEZ/IVA. Weighted UniFrac computed with MetaPhlAn4 R script. PERMANOVA and PERMDISP computed by the vegan package with the functions adonis2 and betadisp, respectively. Ellipse depicts the 95% confidence level. **E)** Principal component analysis of the Bray-Curtis distances between samples with and without recent antibiotic exposure. Distance matrix generated by the R package vegan using the MetaPhlAn4 species table. PERMANOVA and PERMDISP computed in vegan with the functions adonis2 and betadisp, respectively. Ellipse depicts the 95% confidence level.

**Supplementary Figure S3.**
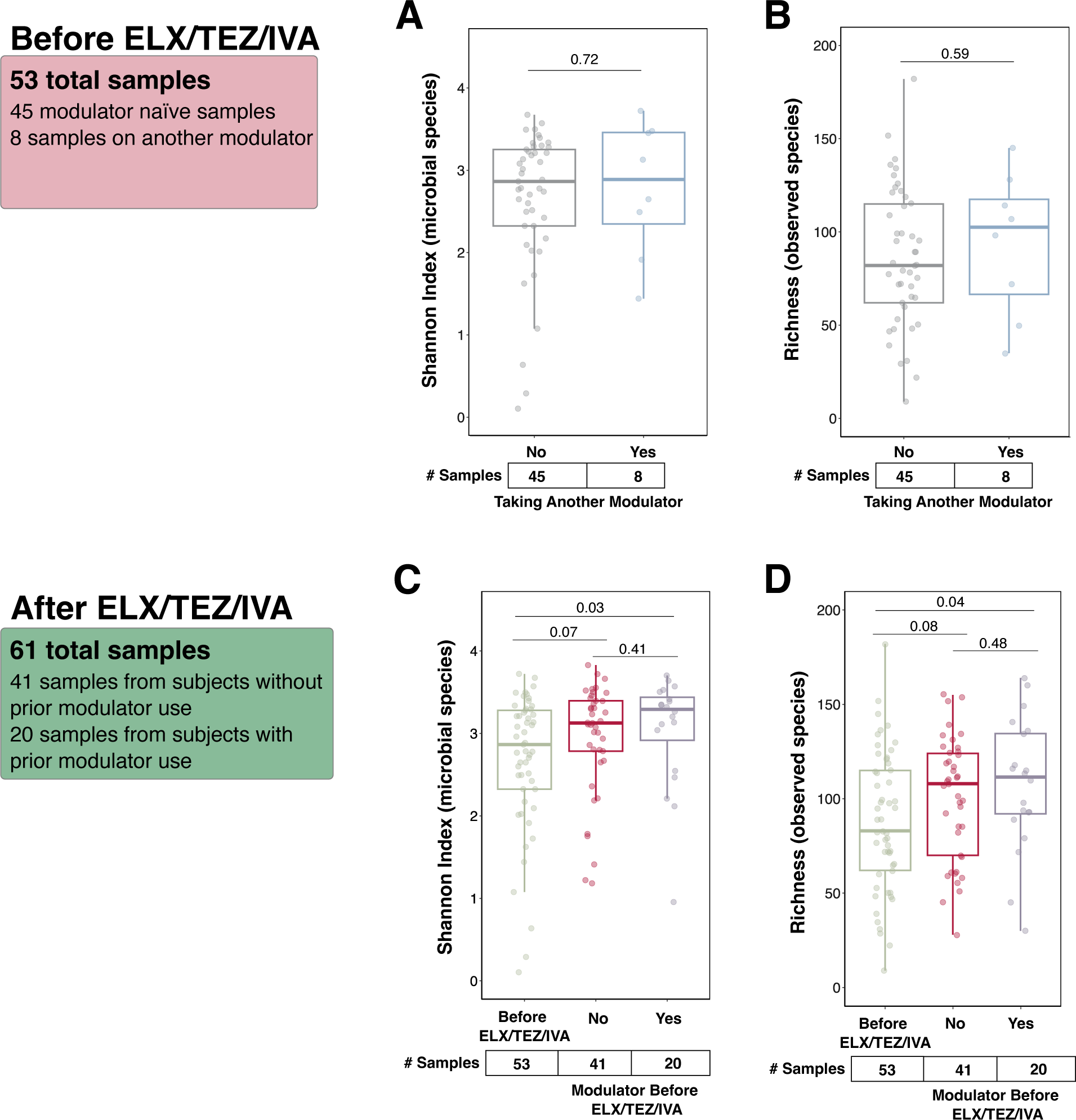
**A-B)** Diversity metrics of pre-ELX/TEZ/IVA samples (samples=53 total). Samples are grouped by whether the subject was receiving another CFTR modulator at the time of sample collection (Yes=8, No=45). **C-D)** Diversity metrics of pre-ELX/TEZ/IVA samples (samples=53) compared to post-ELX/TEZ/IVA samples (samples=61). Post-ELX/TEZ/IVA samples were segregated by whether the subject had previously received any other CFTR modulator (Yes=20 samples, No=41 samples). Shannon Index calculated with the R package vegan using species abundance table from MetaPhlAn4. Microbial richness represents the number of unique species per sample. P values calculated by Wilcoxon rank-sum test.

**Supplementary Figure S4.**
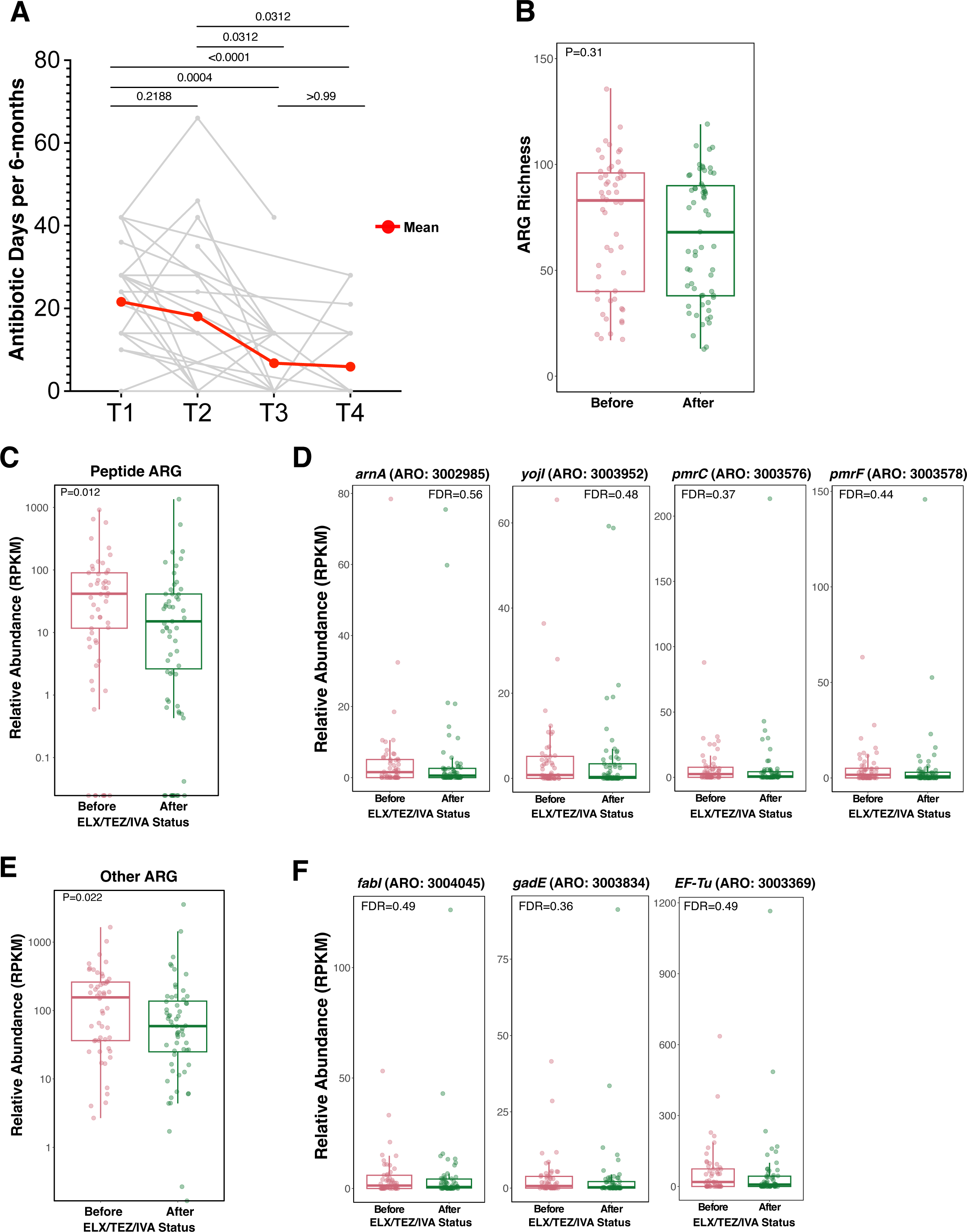
**A)** Cumulative antibiotic days per 6-months prior to stool sample collection. P values calculated by Wilcoxon signed-rank test. **B)** ARG richness (unique genes) before and after ELX/TEZ/IVA. ARGs were profiled using ShortBRED and the Comprehensive Antibiotic Resistance Database (CARD). Each dot represents a stool sample (samples=114). P values calculated by Wilcoxon rank-sum test. **C)** Cumulative relative abundance (RPKM) of ARGs conferring resistance to peptide antibiotics. P values calculated by Wilcoxon rank-sum test. **D)** Relative abundance (RPKM) of the most prevalent ARGs conferring resistance to peptide antibiotics. MaAsLin2 FDR is depicted. **E)** Cumulative relative abundance (RPKM) of ARGs conferring resistance to other antibiotics. P values calculated by Wilcoxon rank-sum test. **F)** Relative abundance (RPKM) of the most prevalent ARGs conferring resistance to other antibiotics. MaAsLin2 FDR is depicted.

**Supplementary Figure S5.**
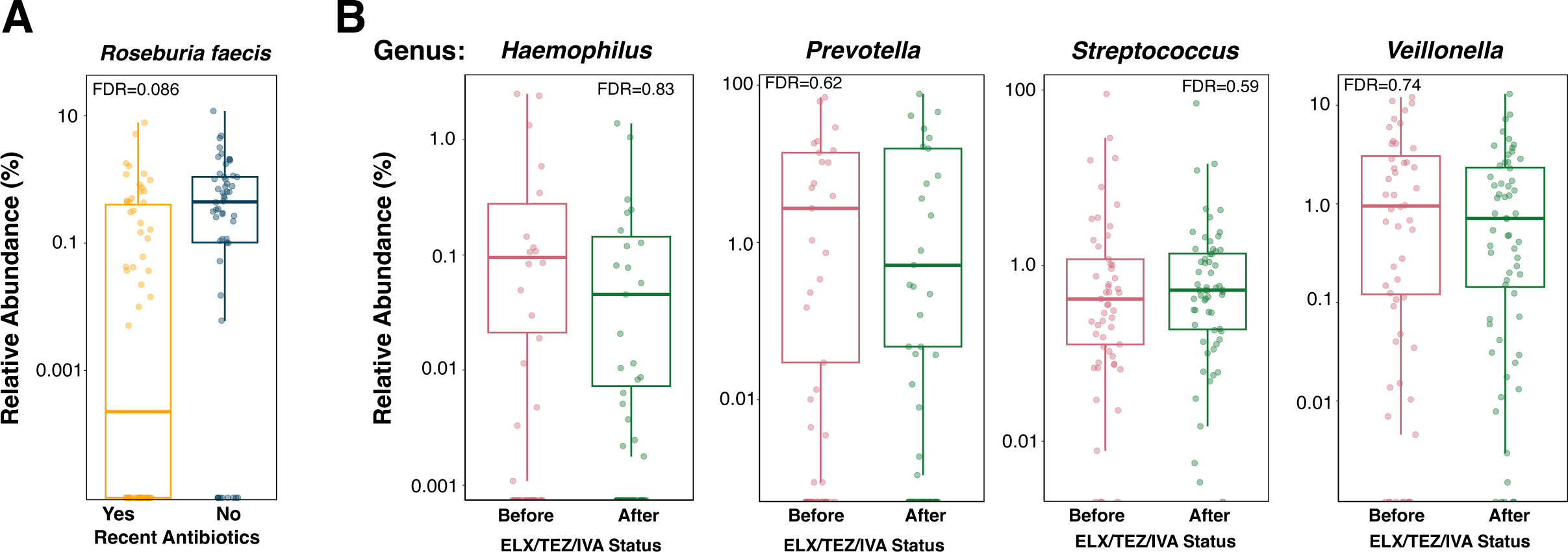
**A)** *Roseburia faecis* relative abundance between samples with and without recent antibiotic exposure. Each dot represents a stool sample (samples=114). MaAsLin2 FDR is depicted. **B)** Relative abundance of respiratory genera in stool samples before and after ELX/TEZ/IVA. Each dot represents a stool sample (samples=114). MaAsLin2 FDR is depicted. Vertical axis is log10 transformed. Zero values are plotted on the horizontal axes.

**Supplementary Figure S6.**
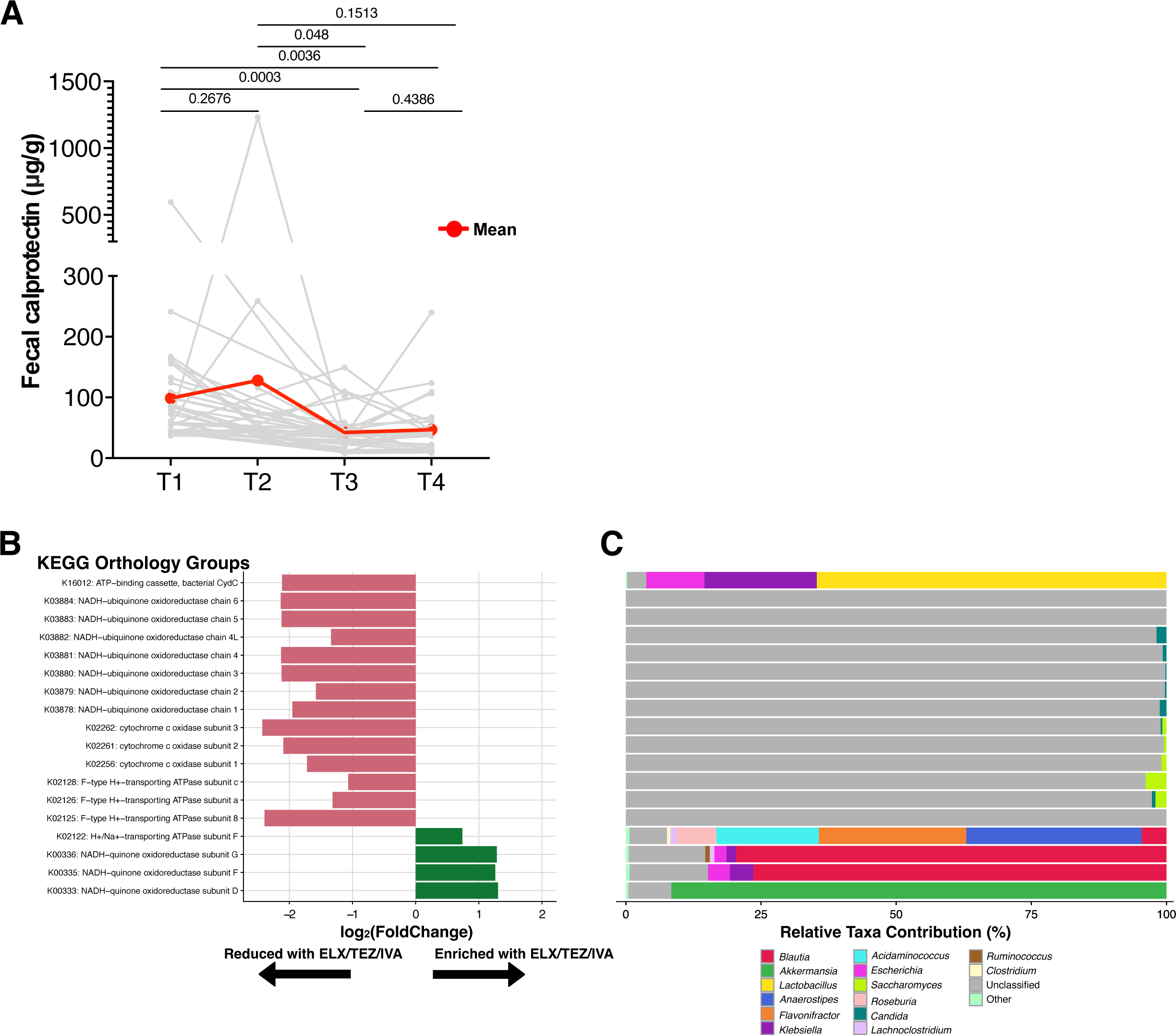
**A)** Fecal calprotectin (μg/g) vs. study timepoint. Red line indicates the mean, and gray lines represent individual patients. P values calculated by Wilcoxon signed-rank test. **B)** Differentially abundant oxidative phosphorylation KEGG orthology (KO) groups from MaAsLin2 (FDR<0.25). Groups correspond to red points in Fig. 5A. log2(FoldChange), which is equivalent to MaAsLin2 coefficient, is depicted. **C)** Taxonomic stratification of differentially abundant oxidative phosphorylation KO groups. KO groups correspond to groups in panel A.

**Supplementary Figure S7.**
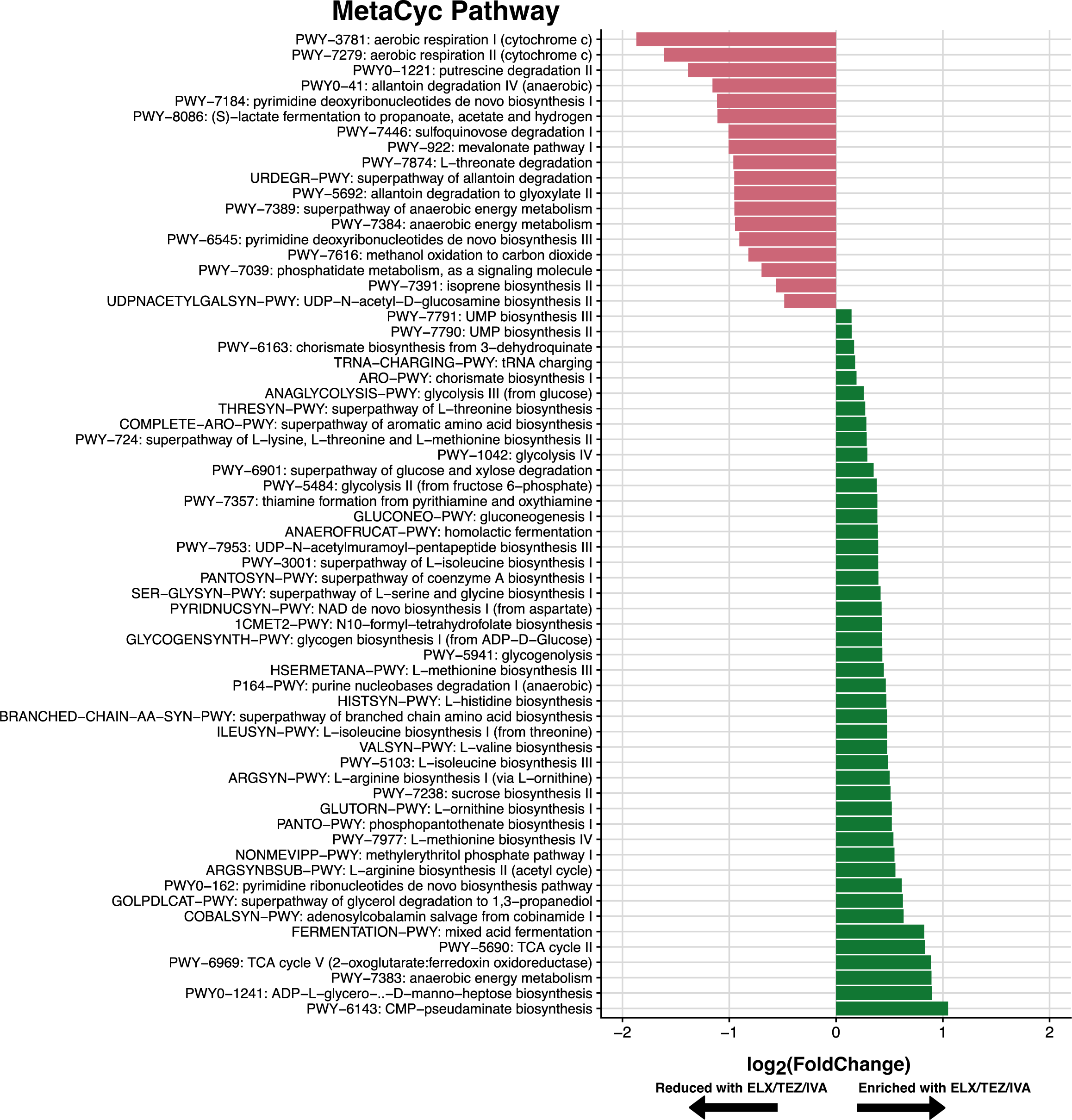
Differentially abundant MetaCyc pathways following MaAsLin2 multivariable association modeling, using ELX/TEZ/IVA status, age, and recent antibiotics as fixed effects in the model. Only pathways with FDR<0.25 are depicted.

## Notes

### Competing Interest Statement

The authors have declared no competing interest.

### Author Declarations

This study was approved by the Monroe Carell Jr. Children's Hospital at Vanderbilt (MCJCHV) Institutional Review Board (IRB # 200396).

### Summary of Updates

Revised in response to reviewer comments. Figures have been rearranged. Discussion section is expanded

